# Effect of Lymphocyte miRNA Expression on Influenza Vaccine-Induced Immunity

**DOI:** 10.1101/2024.11.02.24316654

**Authors:** Iana H. Haralambieva, Tamar Ratishvili, Krista M. Goergen, Diane E. Grill, Whitney L. Simon, Jun Chen, Inna G. Ovsyannikova, Gregory A. Poland, Richard B. Kennedy

## Abstract

Alterations of gene expression by miRNAs contribute substantially to genetic regulation and cellular functions.

We conducted a comprehensive study in 53 individuals before and after the administration of the 2010-2011 seasonal inactivated influenza vaccine to characterize lymphocyte-specific miRNA expression (in purified B cells, CD4+ T cells, CD8+ T cells and NK cells) and its effect on influenza vaccine-induced immune outcomes (hemagglutination inhibition antibody titers/HAI, viral neutralizing antibody titers /VNA and memory B cell ELISPOT).

Overall, we observed relatively stable miRNA expression before/after influenza vaccination. Our statistical analysis uncovered three baseline miRNAs (miR-150-3p, miR-629-5p and miR-4443) that were significantly correlated with influenza vaccine-induced immune outcomes in different cell types. Predictive modeling of influenza vaccine-induced HAI/VNA titers identified a set of specific baseline miRNAs in CD4^+^ T cells as factors predictive of antibody responses. A pathway enrichment analysis on the putative target genes revealed several regulated signaling pathways and functions: TGF-β signaling, PI3K-Akt signaling, p53 signaling, MAPK signaling, TNF signaling and C-type lectin receptor signaling, as well as cell adhesion and adherens junctions, and antiviral host response.

In conclusion, our study offers evidence for the role of epigenetic modification (miRNAs) on influenza vaccine-induced immunity. After validation, identified miRNAs may serve as potential biomarkers of immune response after influenza vaccination.

**Highlights:**

- Host miRNA expression is relatively stable before and after influenza vaccination
- miR-150-3p, miR-629-5p and miR-4443 were correlated with immunity across cell types
- Specific CD4^+^ T cell miRNAs are predictive of antibody responses
- Identified miRNAs may serve as biomarkers of immune response after influenza vaccine

## Introduction

Influenza virus is a pathogen of major public health concern. Seasonal outbreaks can involve millions of cases in the United States and countless more worldwide. Influenza morbidity and mortality is increased in infants, young children, and the elderly. While the impact of influenza on human health has been ameliorated through the use of vaccination, vaccine efficacy rates vary greatly from year to year, indicating a need for the development of improved influenza vaccines, especially in those vulnerable populations. Systems biology studies and high-dimensional platforms are often utilized to comprehensively assess the human transcriptome in order to elucidate how protective immunity develops and is maintained after vaccination and to discover potential biomarkers of protective immunity.

MicroRNAs (miRNAs) are 21-23 nucleotide sequences that modulate gene expression by binding to the 3’ UTR of target mRNAs [1]. They control key biological processes and modulate innate and adaptive antiviral responses. Host miRNAs have also been demonstrated to affect viral gene expression and viral miRNAs have been shown to affect host gene expression [2, 3]. miRNA profiling studies of viral infection have highlighted the important regulatory effect of noncoding RNAs on immune response [2–4].

The goal of the study was to comprehensively assess lymphocyte-specific miRNA expression before and after influenza vaccination and to identify baseline/early miRNA biomarkers associated with, and perhaps predictive of, immune response to influenza vaccination.

## Methods

The methods described here are similar or identical to the ones in our previously published studies [5–8].

### Study subjects

A subgroup of 53 people was randomly selected from a previously recruited cohort of 159 older adults (50-74 years old, males and females) in good general health [7]. Individuals received the 2010-2011 trivalent inactivated influenza vaccine. This vaccine contained the A/California/7/2009 H1N1-like, A/Perth/16/2009 H3N2-like, and B/Brisbane/60/2008-like strains. Blood samples were collected before vaccination (Day 0) and at Day 3 and Day 28 post-vaccination. The Mayo Clinic Institutional Review Board approved all study procedures and written informed consent was obtained from each participant.

### Immunologic Testing

Influenza A/California/7/2009/H1N1-specific hemagglutination inhibition (HAI) titers, viral microneutralization assay (VNA) titers, and B cell ELISPOTs were performed using serum and PBMCs from each subject at each timepoint, as previously described [7, 8].

### Cell purification and miRNA isolation

Cryopreserved PBMCs were thawed using standard procedures; stained with fluorescently labeled antibodies to CD3, CD4, CD8, CD19, and CD56 (BD Biosciences; San Jose, CA); and sorted by fluorescence-activated cell sorting using a FACSAria (BD Biosciences; San Jose, CA). CD4+ T cells, CD8+ T cells, CD3-CD56+ NK cells, and CD19+ B cells were sorted into cold PBS containing 2% fetal bovine serum. Total RNA, including miRNA, was extracted using miRNeasy Mini Kits (Qiagen; Valencia, CA) according to the manufacturer’s instructions.

### Next Generation Sequencing

Extracted miRNA quantity and quality were assessed using an Agilent 2100 Bioanalyzer (Agilent; Palo Alto, CA). Illumina TruSeq miRNA library construction and single-end read sequencing on Illumina HiSeq 2000 (Illumina; San Diego, CA) was performed at the Mayo Clinic Advanced Genomics Technology Center as previously described [5, 9, 10] with twelve samples multiplexed per lane. Subjects were randomized prior to library preparation and three subjects, each with four cell types, were allocated to a lane; hence, all lanes were balanced by cell type.

### Bioinformatics

The bioinformatics pipeline has been previously described [5]. The processing of the FASTQ files was done using a comprehensive pre-processing and analytical pipeline (CAP-miRNA) [11] that assesses read quality (FastQC); trims adapter sequences (Cutadapt); and aligns reads for Integrative Genomics Viewer (IGV) viewing and detection of mature, precursor, and novel miRNAs (miRDeep2) [12].

### Statistical Analysis

The primary analysis focused on the association between immune response variables (HAI, VNA and B cell ELISPOT) and the expression of each mature miRNA by cell subset (CD4+ T cell, CD8+ T cell, B cell, and NK cell). In addition, analyses were conducted to examine changes over time in mature miRNA expression within cell subsets. For each cell subset, non-expressed miRNAs (median counts of zero at all timepoints) were removed from further analysis for that cell subset. In total, 129 B cell, 127 CD4+ T cell, 105 CD8+ T cell, and 103 NK cell samples were carried forward for normalization and analysis. Normalization was done separately for each cell subset using the trimmed mean of M-values method [13] as implemented in the edgeR package [14] in R. A per cell subset post-normalization filter removed miRNA where the median count at each visit were all less than or equal to 4. All correlation analyses were computed using Spearman’s correlation and the normalized values of miRNA expression data. Changes over time in miRNA were compared using Generalized Estimating Equations (GEE) [15] to account for the within-subject correlation. Per-miRNA negative binomial GEE models were fit in SAS© using miRNA as the response variable and the timepoint as the independent variable. The normalization offset and tagwise dispersion were estimated in edgeR [14]. False discovery rate control (BH procedure) was performed to correct for multiple testing and both p-values and FDR-adjusted p-values or q-values were reported. Multivariable models for each of the immune response variables were created using cross-validated, elastic-net penalized regression models, with the tuning parameter α = 0.9 [16]. Models were developed separately for each immune response outcome (HAI, VNA or B-cell ELISPOT); cell subset; and timepoint. The dependent variable in each model was the immune response outcome at Day 28 minus the response at Day 0 (log2 scale), while the normalized miRNA expression values were the independent variables. In order to reduce signal noise, only 25% of the miRNAs with the highest variability between subjects were considered in each model. Sex was included in the models as a covariate. The R package “multiMiR” [17] was used to identify genes representing validated targets of miRNAs of interest. Identified target genes were used in the “clusterProfiler” R package to identify enriched biological (KEGG) pathways regulated by the miRNAs of interest [18]. Results were reported with gene ratio, which represents the ratio of the input genes that are annotated in a term, and FDRs (q-values) of enrichment.

## Results

### miRNA profiles of lymphocyte subsets

Cell subset- (CD4+ T cell, CD8+ T cell, B cell, and NK cell) specific miRNA expression demonstrated that, overall, miRNA expression was similar across different cell types and relatively stable (for most cell subsets) in response to vaccination. Figure 1A displays the common Day 0 miRNA expression between different cell types (miRNA expression overlap), as well as cell type-specific miRNA expression. Supplementary Fig.1 illustrates a heatmap clustering of miRNA expression by cell subset. 683 miRNAs had median counts > 0. Of those 412 miRNAs with a median count > 4 (post-normalization filter), CD4^+^ T cells expressed 358, CD8^+^ T cells expressed 324, NK cells expressed 321, and B cells expressed 301. 62.6% of these miRNAs were expressed in all four lymphocytes, while cell type-specific miRNAs were relatively rare (Fig. 1A). Our initial hypothesis was that shared miRNAs would be involved primarily in cytoskeletal activity, cell cycle control, proliferation/apoptosis, and other housekeeping activities, while cell subset-specific miRNAs (Table 1) would reflect each lymphocyte’s unique immunologic function. Pathway analysis of the predicted targets of cell subset-specific miRNAs, however, indicated that some of the identified pathways were also overlapping consisting of miRNAs predicted to influence general signaling pathways and transcriptional programs involved in T cell, NK cell and B cell homeostasis, development, differentiation, trafficking and functional activity: MAPK signaling, p53 signaling, adhesion and adherens junctions, cellular senescence, as well as in macromolecule biosynthesis, metabolism, cell-to-cell interactions, endocytosis, cytoskeleton regulation, and anti-viral pathways (Supplementary Fig. 2). Importantly, the analysis identified miRNA target pathways predicted to influence B and T cell receptor signaling and effector functions: MAPK, mTOR, PI3K-Akt, Ras signaling (Supplementary Fig. 2).

**Fig. 1.**
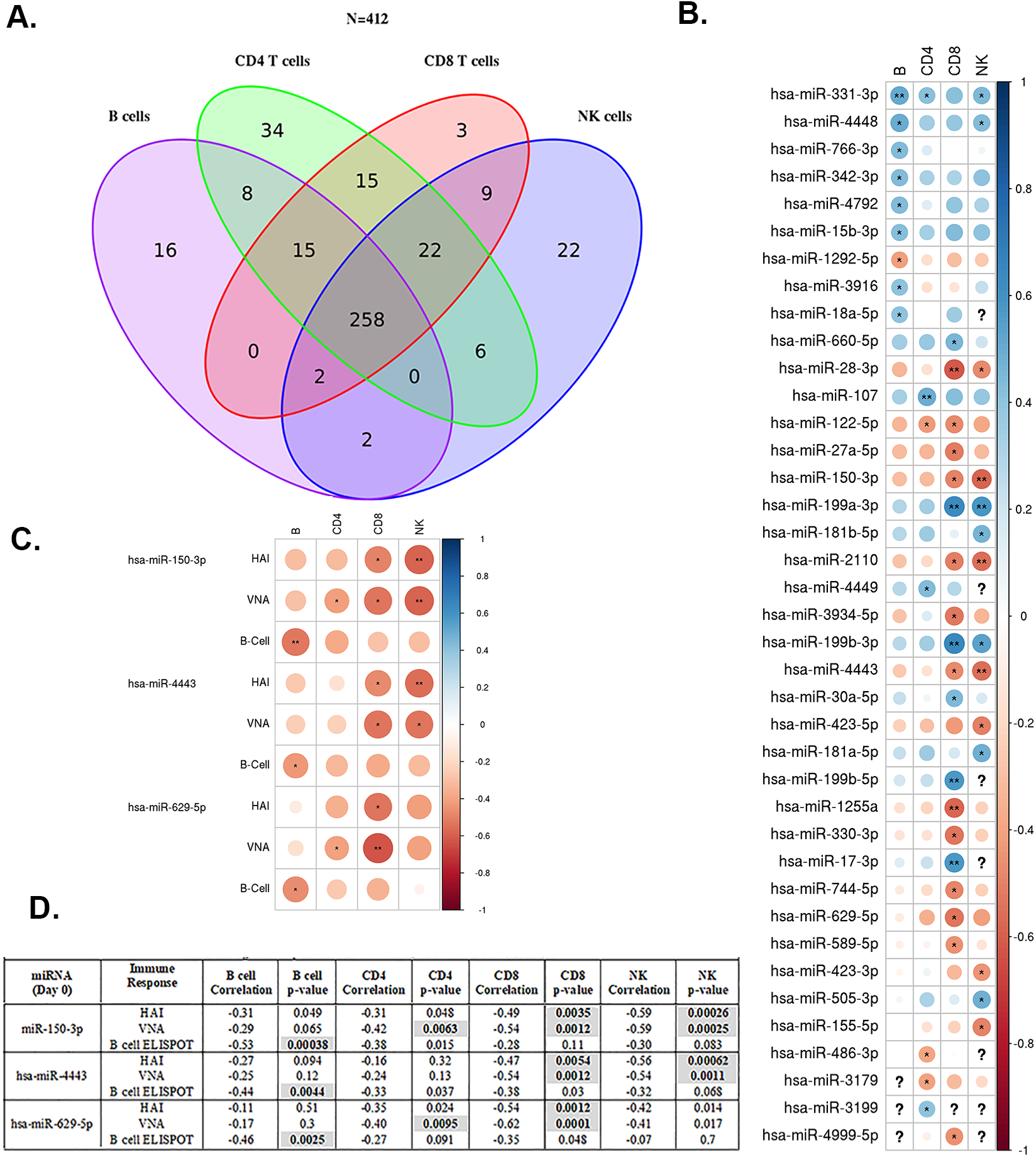
Baseline miRNA expression and its influence on humoral immune response outcomes following influenza vaccination **A.** Venn diagram of miRNAs expressed within each cell type**. B.** Correlation between baseline miRNA expression and HAI antibody titer following influenza vaccination. Correlation plot illustrating the associations between baseline miRNA and HAI antibody titer (Day 28 -Day0) for each cell subset (CD4+ T cell, CD8+ T cell, B cell, and NK cell) **C.** Baseline miRNAs significantly correlated with all humoral immune outcomes (change in HAI antibody titer, VNA antibody titer and B cell ELISPOT response) for each cell subset. The color of the correlation represents the direction (red, negatively associated, blue positively associated with the immune outcome). The size of the circle represents the strength of the correlation. The color intensity scale to the right of the figure is a scale for the Spearman’s correlations with darker colors indicating stronger correlation. Blank cells indicate that the correlation with the specific miRNA was zero in the associated cell type, and cells with a “?” indicate that the miRNA was not present (not measured) in the cell type. *p <0.01, **p<0.001. **D.** miRNAs significantly correlated with HAI, VNA, and B cell ELISPOT. Correlations are with the immune response deltas (Day28 – Day0). P<0.01 was used as the cut-off.

**Table 1.** Cell type-specific miRNAs.

| **Table 1. Cell Type-Specific miRNAs** |  |  |  |
| --- | --- | --- | --- |
| **B cells (n=16)** | **NK cells (n=22)** | **CD4 T cells (n=34)** | **CD8 T cells (n=3)** |
| hsa-miR-1295a | hsa-miR-10a-3p | hsa-let-7e-3p | hsa-miR-199a-5p |
| hsa-miR-129-5p | hsa-miR-1908 | hsa-let-7g-3p | hsa-miR-584-5p |
| hsa-miR-1303 | hsa-miR-196b-5p | hsa-miR-1254 | hsa-miR-9-3p |
| hsa-miR-138-5p | hsa-miR-20b-3p | hsa-miR-125b-2-3p |  |
| hsa-miR-195-3p | hsa-miR-2355-3p | hsa-miR-1261 |  |
| hsa-miR-195-5p | hsa-miR-338-3p | hsa-miR-1285-5p |  |
| hsa-miR-3150b-3p | hsa-miR-429 | hsa-miR-1299 |  |
| hsa-miR-3614-5p | hsa-miR-432-5p | hsa-miR-1976 |  |
| hsa-miR-4491 | hsa-miR-4489 | hsa-miR-296-5p |  |
| hsa-miR-5196-3p | hsa-miR-4665-5p | hsa-miR-34a-5p |  |
| hsa-miR-574-3p | hsa-miR-4755-5p | hsa-miR-365a-3p |  |
| hsa-miR-577 | hsa-miR-4772-3p | hsa-miR-365b-3p |  |
| hsa-miR-582-3p | hsa-miR-4772-5p | hsa-miR-3687 |  |
| hsa-miR-629-3p | hsa-miR-550a-3-5p | hsa-miR-374b-3p |  |
| hsa-miR-708-3p | hsa-miR-550a-5p | hsa-miR-4536-5p |  |
| hsa-miR-708-5p | hsa-miR-6131 | hsa-miR-455-5p |  |
|  | hsa-miR-628-5p | hsa-miR-4688 |  |
|  | hsa-miR-641 | hsa-miR-4781-3p |  |
|  | hsa-miR-873-3p | hsa-miR-4791 |  |
|  | hsa-miR-876-3p | hsa-miR-506-3p |  |
|  | hsa-miR-92a-2-5p | hsa-miR-508-3p |  |
|  | hsa-miR-93-3p | hsa-miR-508-5p |  |
|  |  | hsa-miR-5090 |  |
|  |  | hsa-miR-509-3-5p |  |
|  |  | hsa-miR-509-3p |  |
|  |  | hsa-miR-513c-5p |  |
|  |  | hsa-miR-514a-3p |  |
|  |  | hsa-miR-548b-5p |  |
|  |  | hsa-miR-548q |  |
|  |  | hsa-miR-6511a-3p |  |
|  |  | hsa-miR-6511b-3p |  |
|  |  | hsa-miR-659-5p |  |
|  |  | hsa-miR-676-3p |  |
|  |  | hsa-miR-939-5p |  |

miRNAs with a median count of less than or equal to 4 were not included.

We also examined the 20 miRNAs with the highest median baseline expression level in each cell type and found that the highly expressed miRNAs are largely overlapping (Supplementary Table 1). There were 22 miRNAs that were in the top 20 for at least two cell subsets (Supplementary Table 1).

### miRNA changes over time after vaccination

Overall, miRNA profiles in B cells, CD8+ T cells, and CD4+ T cells were stable and barely changed after influenza vaccination. We did not discover significant expression changes (q<0.1) in CD4+ T cells or B cells. In CD8+ T cells, the expression of only six miRNAs (miR-143-3p, miR-152, miR-196a-5p, miR-548f, miR-9-3p, and miR-9-5p) decreased on Day 28 following influenza vaccination. Interestingly, NK cell miRNA expression was much more dynamic and changes were identified earlier, with 142 miRNAs exhibiting significant expression changes on Day 3 following vaccination (Supplementary Table 2). Pathway analysis of the predicted targets of the top NK cell miRNAs (n=24), indicated the regulation of the following pathways: ErbB signaling pathway (p=2.38E^-06^), Hippo signaling pathway (p=1.98E^-05^), Wnt signaling pathway (p=3.04E^-05^), MAPK signaling pathway (p=7.40E^-05^), Ras signaling pathway (p=0.001) and mTOR signaling pathway (p=0.001) among others (data not shown).

### miRNAs correlated with influenza vaccine response

Since the expression of miRNAs in B and T lymphocytes was stable across timepoints, we first explored if the baseline expression level of multiple miRNAs in each cell type correlated with influenza vaccine-induced (Day 28 – Day 0) HAI response (Fig. 1B, Supplementary Table 3). We identified 39 miRNAs with baseline expression significantly correlated with HAI response (Fig. 1B, p˂0.01). Importantly, for nine of the miRNAs (23% of the identified miRNAs), the response correlations were significant across more than one cell subset (Fig.1B at p˂0.01). For example, the positive correlation between miR-331-3p expression and HAI was significant for B cells, CD4+ T cells, and NK cells (p<0.01, Supplementary Table 3, Fig.1B), while the miR-331-3p correlation in CD8+ T cells was suggestive (p=0.016, Supplementary Table 3). Our results also indicate a large overlap in the HAI-correlated miRNAs between CD8+ T cells and NK cells (6/14 NK miRNAs also found in CD8+ T cells) (Fig. 1B, Supplementary Table 3). Importantly, the baseline expression levels of three miRNAs – miR-150-3p, miR-4443, and miR-629-5p – were significantly correlated with all three immune outcomes (HAI, VNA, B cell ELISPOT) in one or more cell subsets (Fig. 1C and D), which indicates the importance of these miRNAs in the regulation of influenza-specific humoral immunity. miRNAs correlated with VNA and memory B cell ELISPOT responses are listed in Supplementary Table 3, respectively. Of note, the same 14 miRNAs correlated with B cell ELISPOT response in CD4+ T cells were also correlated with B cell ELISPOT response in the CD8+ T cell samples.

### Modeling of influenza vaccine-induced immune outcomes

It is expected that multiple miRNAs work in concert to regulate immune response. Thus, a multivariable model, which jointly models the miRNAs in relation to the immune outcomes, could be a more powerful approach to identify predictive miRNAs compared to the “per miRNA” correlation analysis. Therefore, we next applied elastic net regularized linear regression modeling of immune outcomes as a function of miRNA expression. This joint modeling revealed several sets of miRNAs expressed in the different cell subsets that were collectively associated with and predictive of influenza vaccine-induced response outcomes. Figure 2 illustrates the correlation coefficients for the individual miRNAs baseline expression associated with each model. The top two models from our analysis efforts refer to baseline miRNA expression in CD4^+^ T cells correlated with both HAI (Fig. 2A) and VNA (Fig. 2B) responses, with extensive overlap in the identified miRNAs. Similarly, baseline miRNA expression in CD8+ and B cells correlated with VNA response and both models contained miR-4792 and miR-331-3p (Fig. 2C and D). For the two top models (Fig. 2A and B), we performed pathway enrichment analysis (as outlined in Statistical Analysis) on the putative target genes. These results are summarized in Fig. 3 and represent evidence for CD4+ T cell miRNA influence on the following biological processes and signaling pathways impacting immune response: carbohydrate and lipid metabolism, endocytosis and cytoskeleton regulation, cell adhesion and adherens junctions, TGF-β signaling, MAPK signaling, p53 signaling, PI3K-Akt signaling and C-type lectin receptor signaling pathways among others. Our predictive modeling efforts of immune outcomes as a function of miRNAs post-vaccination revealed only a small number of CD4^+^ T cell/NK cell miRNAs correlated with influenza-specific memory B cell ELISPOT response or HAI titer (Supplementary Fig. 3A, B and C).

**Fig. 2.**
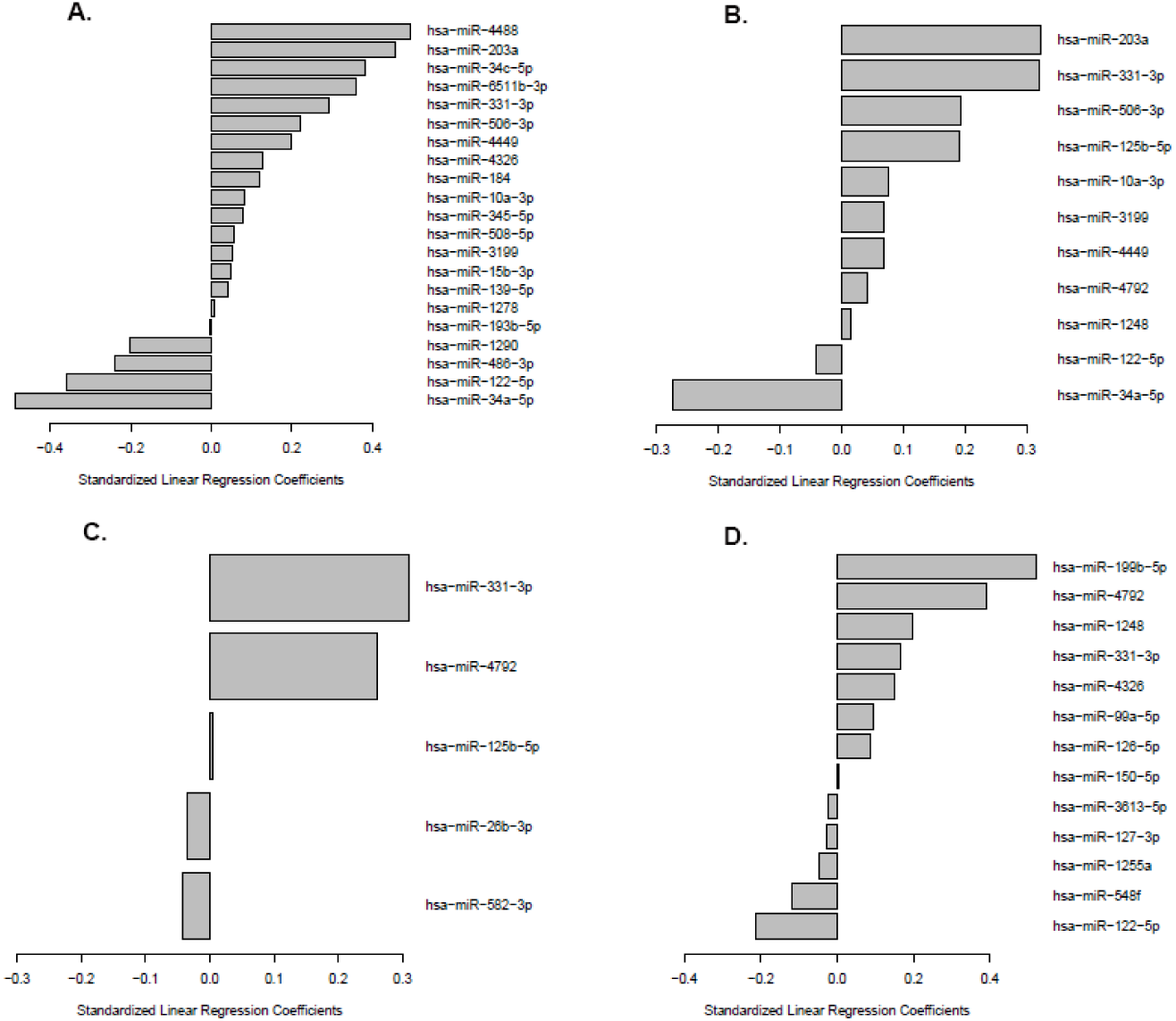
Predictive modeling of baseline miRNA expression on influenza vaccine-induced humoral immune outcomes. Modeling was performed as described in Statistical Analysis. The baseline CD4+ T cell miRNAs predictive of influenza vaccine-induced HAI titer are depicted in **A**. and those predictive of VNA titer are depicted in **B**. The baseline B cell miRNAs and CD8+ T cell miRNAs predictive of influenza vaccine-induced VNA titer are depicted in **C** and **D**, respectively. The standardized linear regression coefficients are the estimated change in the response for a one standard deviation change in the associated miRNA expression. Positive coefficient values (bars extending to the right of the centerpoint of the x-axis) indicate that an increase in the miRNA expression is associated with an increase in the immune response outcome, while negative coefficient values (bars extending to the left of the centerpoint of the x-axis) indicate that as the miRNA expression increases the immune. response outcome decreases.

**Fig. 3.**
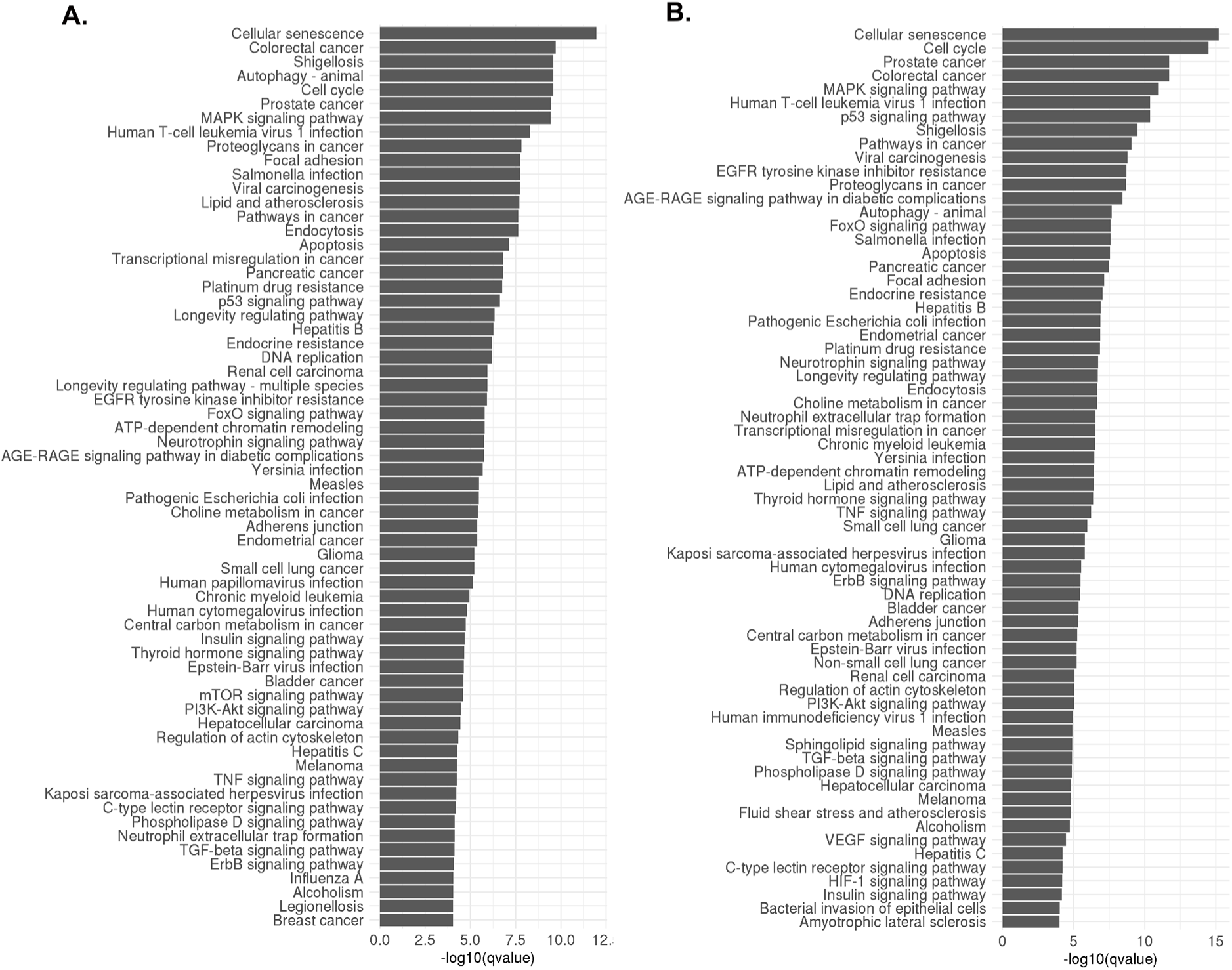
Pathway analysis of target genes regulated by baseline CD4+ T cell-specific miRNAs influencing influenza vaccine-induced HAI/VNA antibody titer. Enrichment pathway analysis performed (see. Statistical Analysis) on the predicted gene targets of baseline CD4+ T cell miRNAs associated with **A.** HAI titer or **B.** with VNA titer. The bar length corresponds to the significance level of enrichment. Only pathways with q<0.0001 are included in the graph.

## Discussion

Our study assessed miRNA expression before/after influenza vaccination in four major lymphocyte cell types (CD4+ T cell, CD8+ T cell, B cell, and NK cell) and demonstrated stable miRNA expression profiles over time (with the exception of NK cell miRNAs) and noticeably overlapping miRNAs expressed across different cell types. The interrogation of lymphocyte subset-specific miRNAs at baseline revealed miRNA control of both common and unique functional pathways. Among the targeted unique pathways are pathways closely involved in functional activity of specific cell types. These include the TGF-beta signaling pathway, apoptosis and the related to cell cycle control/longevity FoxO signaling pathway in CD4+ T cells; the PI3K-Akt signaling pathway important for the B cell development/differentiation in B cells; and the Ras signaling cascade, triggered downstream of B cell receptor to regulate B cell survival and function. miRNAs also regulated relatively common pathways linked to metabolism and proliferation, cellular senescence, cell adhesion-related pathways, MAPK signaling, p53 signaling, as well as endocytosis, cytoskeleton regulation, and anti-viral pathways. It is likely that the common pathways reflect the cellular housekeeping activity, cell cycle progression, cytoskeletal functions and metabolism necessary to support lymphocyte activation, differentiation, proliferation, and functional/immune activity.

Although we expected to find more pronounced changes in cellular miRNA expression levels - across timepoints relative to vaccination (baseline, Day 3, Day 28), we found that miRNAs in B cells, CD4+ T cells, and CD8+T cells were relatively stable over time following vaccination. Possible explanations of this observation include: sub-optimal timing of miRNA expression assessment following vaccination (i.e., miRNA expression changes were detectable at other timepoints); miRNA expression changes existed in antigen-specific cells and were missed with global miRNA expression assessment; or influenza vaccination did not alter significantly the host miRNA expression landscape. Interestingly, NK cells exhibited significant changes in miRNA expression at Day 3 post-vaccination relative to baseline. Several of these NK miRNAs have been implicated in biological activities that impact NK cell activity. Expression levels of miR152 changed significantly on Day 3 in NK cells, as well as on Day 28 in CD8+ T cells. miR152 has been demonstrated to control HLA-G expression and NK cell-mediated cytolytic activity [19].

Another interesting finding, miR-125b-5p, has been reported to inhibit IRF4, which leads to increased expression of MICA a ligand for the NK cell activating receptor NKG2D [20]. Downregulation of this molecule after vaccination would promote enhanced NK cell activity. NK cells are important innate lymphocytes that have innate immune memory and are able to significantly regulate adaptive immune responses after vaccination [21]. For example, NK cell cytolytic activity was demonstrated to affect antigen persistence and/or target responding T cells after vaccination [21, 22]. Other NK cell functional activities (secretion of IFNγ and expression and activating/inhibitory receptors) have been linked to the stimulation of antigen-presenting cells following vaccination [23]. This suggests that changes in NK cell miRNA expression following vaccination can have direct or indirect effect on adaptive immune responses after influenza vaccination. Other miRNAs exhibiting NK cell expression changes include miR126-5p, which has been implicated in the macrophage response to LPS [24] and in endothelial cell angiogenesis, [25] and miR148a-3p, which in concert with other miRNAs has been demonstrated to modulate plasma cell differentiation in B cells [26]. miR-339-5p has been shown to regulate lymphocyte tissue migration [27, 28] and to control inflammation by inhibiting IKK-β and IKK-ε activity [29]. Further investigation will be necessary in order to understand what role, if any, these NK cell miRNAs play in cell activation, proliferation, or immune function.

Of great interest to vaccinology is the discovery of miRNAs that may regulate immune response and/or potentially serve as biomarkers of vaccine-induced immunity for inclusion in an immune signature predictive of optimal (or suboptimal) vaccine response. In this study, three baseline miRNAs (miR-150-3p, miR-629-5p and miR-4443) were significantly negatively correlated with all three immune outcomes (HAI titer, VNA titer and memory B cell ELISPOT response) when data from all four cell types were analyzed (Fig. 1C and D). miR-150-3p has previously been reported to regulate key biological processes such as inflammation, apoptotic pathways and cell proliferation [30]. miR-150 overexpression has been found to profoundly impair formation of mature B cells and their functional activity with smaller effect on the development of T cells, granulocytes and macrophages [31]. Similarly, in our study, increased miR-150 expression in B cells was negatively correlated (p=0.00038) with memory B cell ELISPOT response after influenza vaccine, likely due to mature B cell suppression/suppression of germinal center memory B cell differentiation [32], while the increased expression in T cells (CD4+ and CD8+ T cells) was negatively correlated with the HAI/VNA titer. It was previously established that the intracellular miR-150 expression in T cells (e.g., CD4+ T helper cells) is downregulated upon T cell activation to control key genes underlying T cell functions due to its quick extracellular release in exosomes [33, 34]. Importantly, extracellular miR-150 level (corresponding to the level of downregulation of intracellular miR-150 in T cells) has been established as a reliable serum marker of lymphocyte activation (T cell activation) [35]. Moreover, the serum level of miR-150 was found to be elevated in subjects following MF-59-adjuvanted 2009 influenza A (H1N1) vaccination with higher levels observed in subjects with higher antibody titers [33]. This is corroborated by our finding of negative correlation between the intracellular expression of miR-150 in T cells and influenza vaccine-induced HAI/VNA antibody responses. Thus, miR-150 expression emerges as a reliable biomarker of influenza vaccine-induced immunity as identified in a previous study [33] and ascertained by our findings. miR-629 has been investigated as a prognostic marker in cancer and has been implicated primarily in the regulation of proliferation, apoptosis, cell migration and alteration of TGF-beta/Smad signaling (via its target gene TRIM33) [36, 37]. Its expression has not been previously linked to immune response to vaccination/infection as suggested by our findings. miR-4443 has been connected to the regulation of key cellular signaling pathways such as the NF-κB, TGF-β and the Ras signaling pathways [38]. Increased expression of miR-4443 has been linked to dysfunction in T cells mediated via TNF receptor associated factor 4/TRAF4, and thus it is possible that this miRNA negatively impacts humoral immunity by impairing CD4+ T cell function [39]. On the other hand, the Ras GTPases serve as signaling checkpoints in antigen receptors, and Ras signaling (regulated in part by the expression of miR-4443) may play a key role in immune cell activation and function by engaging multiple downstream effectors [40, 41]. The Ras signaling pathway downstream of BCR is known to be a critical part of B cell proliferation, differentiation, antigenic activation and functional activity [42]. The Ras-ERK1/2 MAPK cascade is also tightly involved in Th2 cell differentiation [43], so it is possible that miRNA expression (miR-4443 and other miRNAs) regulating these signaling pathways may fine-tune critical antigen-driven cellular activities in B and T lymphocytes.

Other miRNAs of interest demonstrated significant associations with HAI antibody titer (and not other immune outcomes) after influenza vaccination, but across several cell subsets. For example, the baseline expression of miR-331-3p was positively associated with HAI in B cells, CD4+ T cells, and NK cells (Fig. 2).

Our predictive modeling efforts of baseline miRNA expression confirmed that indeed miR-331-3p is an important biomarker of influenza vaccine-induced immunity as it was identified as a positive predictor of HAI/VNA antibody titer in CD4+ T cells, B cells and CD8+ T cells (Fig. 2). miR-331-3p has been demonstrated to inhibit proliferation and support apoptosis via regulating the PI3K/Akt and ERK1/2 pathways [44, 45]. It is possible that miR-331-3p expression regulates cell proliferation in several immune cell types and thus contributes to their functional activities following vaccination. For example, both signaling pathways (altered by the expression of miR-331-3p) are key in regulating B cell metabolic activity, proliferation and differentiation upon antigenic stimulation and thus are likely to influence vaccine-induced immunity [46]. miRNAs of the miR-17–92 locus (i.e., miR-17 and miR-18a) in B and T cells (Fig.2) were also identified as being associated with post-vaccine HAI titer. These miRNAs play an important role in B cell development, differentiation, survival and regulation of class switching, and thus are intimately involved in B cell functionality [47]. The baseline cell expression of miR-15 and miR-223 was found to be associated with post-vaccine HAI titer and/or influenza virus-specific memory B cell ELISPOT response in our study. miR-15 was previously demonstrated to target Bcl2 and to regulate B cell survival, while miR-223 is enriched in the memory B cell compartment and regulates an important transcription factor (LMO2) critical for B cell differentiation, and has been also established as a regulator of activation-induced cytidine deaminase (AID) [48]. AID is an essential enzyme mainly expressed in germinal center B cells that plays a role in somatic hypermutation/antibody diversity and Ig class switching [49]. Thus, both miR-15 and miR-223 emerge as plausible regulators of B cell function and biomarkers of influenza. vaccine-induced humoral immunity. The expression of miR-34a-5p in CD4+ T cells was identified as a plausible predictor of influenza vaccine-induced antibody response (Fig. 2). The involvement of miR-34a-5p in plasma cell differentiation (along with miR-125b-5p and miR-101-3p) has been previously demonstrated [26], but its role in regulating T cell functionality is still unclear. Interestingly, the expression of miR-125b-5p has also been identified as a predictor of VNA antibody response in both B cells and CD4+ T cells in our study.

The top predictive models from our study (Fig. 2A and B) identified specific baseline miRNAs in CD4^+^ T cells as predictors (or factors associated with) of influenza vaccine-induced HAI/VNA antibody responses, with some overlap in the identified miRNAs (for the HAI and the VNA model). Interestingly, models involving miRNA expression post-vaccination resulted in less robust findings (Supplementary Fig. 3). The pathway enrichment analysis on the putative target genes regulated by the identified baseline miRNAs revealed the impact of several important signaling pathways in CD4+ T cells in shaping the influenza vaccine-induced immunity. These include TGF-β signaling, PI3K-Akt signaling, MAPK signaling, p53 signaling, TNF signaling and C-type lectin receptor signaling pathways. Basic cellular activities and functions (regulated by these miRNAs) were also identified as important and impacting influenza vaccine-induced humoral immunity: cell adhesion and adherens junctions, antiviral host response, carbohydrate and lipid metabolism, endocytosis and cytoskeleton regulation.

The strengths of our study include the miRNA expression assessment in 4 purified cell subsets (B cells, CD4+ T cells, CD8+ T cells and NK cells) of subjects before/after influenza vaccination and the robust statistical modeling of influenza vaccine-induced immunity. The limitation is the lack of replication of the identified miRNA biomarkers/pathways in an independent cohort. Important next steps will be experimental validation of the identified miRNAs: co-expression studies of miRNAs and putative targets to provide experimental proof of specific miRNA-mRNA interactions; and experiments revealing how miRNAs modulate target protein expression and how this impacts immune response [50].

In conclusion, our study uncovered novel information regarding the complex genetic control of immune pathways, leading to better understanding of immune function regulation. The study also identified plausible predictive biomarkers of influenza vaccine-induced immune response outcomes, that warrant validation in future studies.

## Data availability

All data is available at ImmPort.org (SDY67).

## Acknowledgements

We would like to thank the subjects who volunteered to participate in our study. The study was funded by the National Institute of Allergic and Infectious Diseases of the National Institutes of Health (U01 AI 089859/Bioinformatics Approach to Influenza A/H1N1 Vaccine Immune Profiling and R01AI132348), the Human Immunology Project Consortium Infrastructure and Opportunities Fund, and a grant from the Mayo Clinic Biomarker Discovery program (grant C4330915).

The content is solely the responsibility of the authors and does not necessarily represent the official views of the National Institutes of Health.

