## Supplemental Table 1-3 for "Effect of Lymphocyte miRNA Expression on Influenza Vaccine-Induced Immunity"

**Supplementary Material:**

**Supplementary Fig. 1 Heatmap of miRNA expression by cell subset**

Heatmap of miRNA expression in specific cell subsets. The color gradient from blue to white (to the right) indicates miRNA expression level. The color intensity represents the log-base 2 normalized miRNA expression values, while the white space indicates that the miRNA was not expressed. Samples are arranged in columns by cell subsets indicated by color bars. Raws represent miRNAs clustered using complete link hierarchical clustering with euclidean distance.

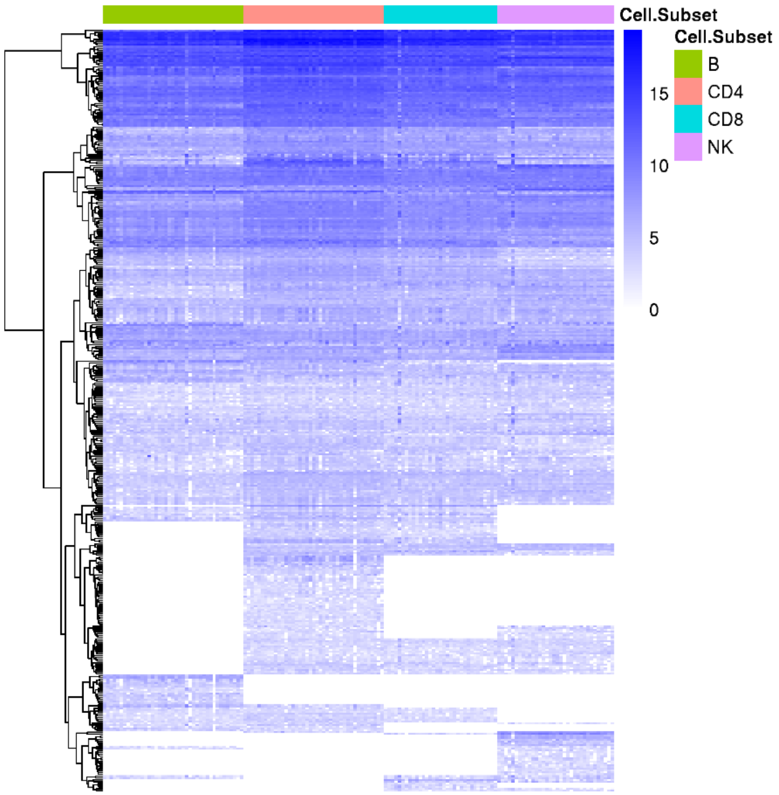
Cell.Subset
Cell.Subset
B
CD4
CD8
NK
15
10
5
0

**Supplementary Fig. 2 Pathway analysis of target genes regulated by cell subset-specific miRNAs expressed at baseline**

Enrichment pathway analysis performed (see. Statistical Analysis) on the predicted gene targets of baseline cell subset-specific miRNAs in **A.** B cells, **B.** CD4+ T cells, **C.** CD8+ T cells and **D.** NK cells. The bar length corresponds to the significance level of enrichment. Only pathways with $q<0.0001$ are included in the graph.

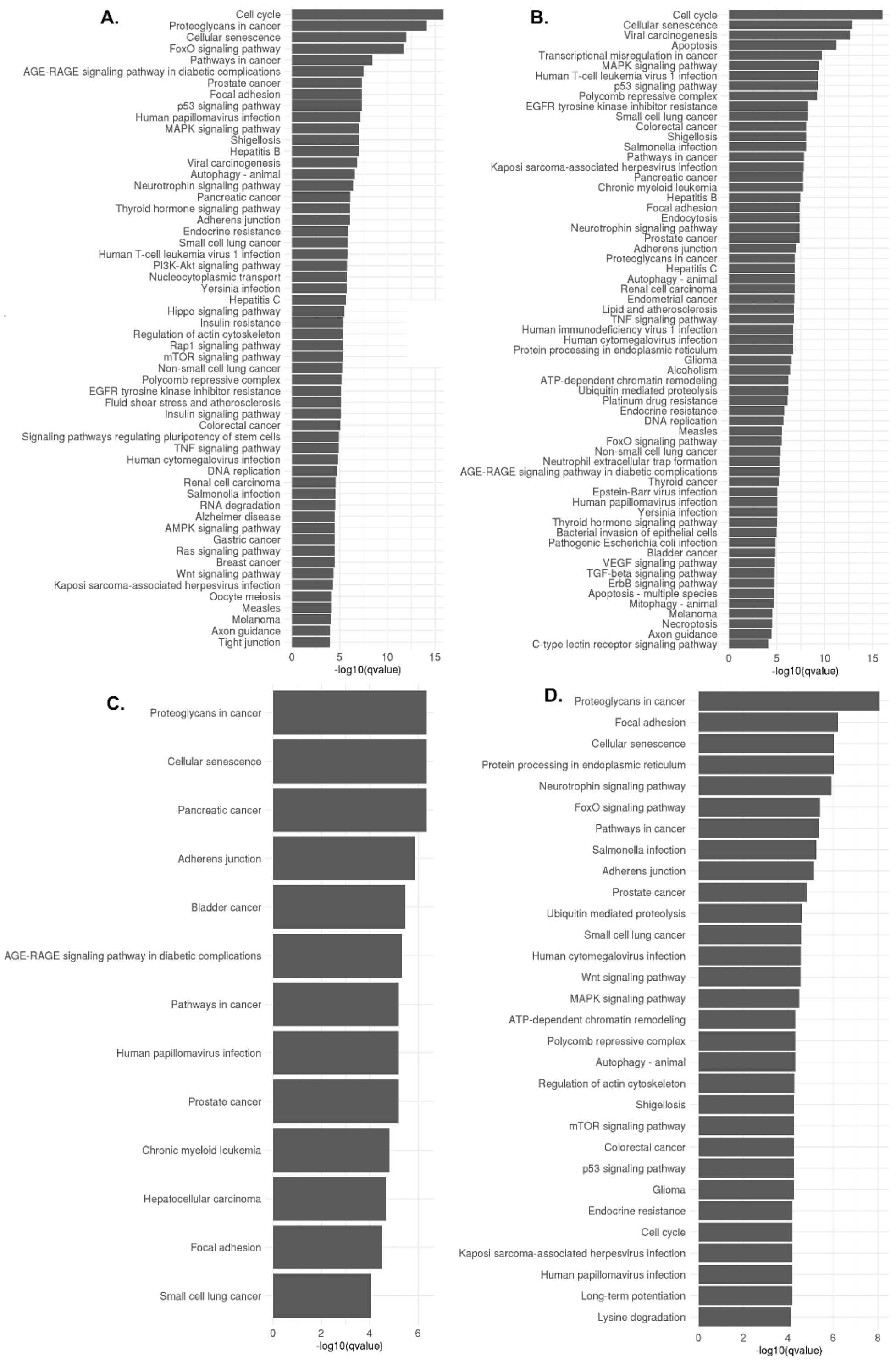
A.
Cell cycle
Proteoglycans in cancer
Cellular senescence
FoxO signaling pathway
Pathways in cancer
AGE-RAGE signaling pathway in diabetic complications
Prostate cancer
Focal adhesion
p53 signaling pathway
Human papillomavirus infection
MAPK signaling pathway
Shigellosis
Hepatitis B
Viral carcinogenesis
Autophagy - animal
Neurotrophin signaling pathway
Pancreatic cancer
Thyroid hormone signaling pathway
Adherens junction
Endocrine resistance
Small cell lung cancer
Human T-cell leukemia virus 1 infection
PI3K-Akt signaling pathway
Nucleocytoplasmic transport
Yersinia infection
Hepatitis C
Hippo signaling pathway
Insulin resistance
Regulation of actin cytoskeleton
Rap1 signaling pathway
mTOR signaling pathway
Non-small cell lung cancer
Polycomb repressive complex
EGFR tyrosine kinase inhibitor resistance
Fluid shear stress and atherosclerosis
Insulin signaling pathway
Colorectal cancer
Signaling pathways regulating pluripotency of stem cells
TNF signaling pathway
Human cytomegalovirus infection
DNA replication
Renal cell carcinoma
Salmonella infection
RNA degradation
Alzheimer disease
AMPK signaling pathway
Gastric cancer
Ras signaling pathway
Breast cancer
Wnt signaling pathway
Kaposi sarcoma-associated herpesvirus infection
Oocyte meiosis
Measles
Melanoma
Axon guidance
Tight junction
0
5
10
15
-log10(qvalue)
B.
Cell cycle
Cellular senescence
Viral carcinogenesis
Apoptosis
Transcriptional misregulation in cancer
MAPK signaling pathway
Human T-cell leukemia virus 1 infection
p53 signaling pathway
Polycomb repressive complex
EGFR tyrosine kinase inhibitor resistance
Small cell lung cancer
Colorectal cancer
Shigellosis
Salmonella infection
Pathways in cancer
Kaposi sarcoma-associated herpesvirus infection
Pancreatic cancer
Chronic myeloid leukemia
Hepatitis B
Focal adhesion
Endocytosis
Neurotrophin signaling pathway
Prostate cancer
Adherens junction
Proteoglycans in cancer
Hepatitis C
Autophagy - animal
Renal cell carcinoma
Endometrial cancer
Lipid and atherosclerosis
TNF signaling pathway
Human immunodeficiency virus 1 infection
Human cytomegalovirus infection
Protein processing in endoplasmic reticulum
Glioma
Alcoholism
ATP-dependent chromatin remodeling
Ubiquitin mediated proteolysis
Platinum drug resistance
Endocrine resistance
DNA replication
Measles
FoxO signaling pathway
Non-small cell lung cancer
Neutrophil extracellular trap formation
AGE-RAGE signaling pathway in diabetic complications
Thyroid cancer
Epstein-Barr virus infection
Human papillomavirus infection
Yersinia infection
Thyroid hormone signaling pathway
Bacterial invasion of epithelial cells
Pathogenic Escherichia coli infection
Bladder cancer
VEGF signaling pathway
TGF-beta signaling pathway
ErbB signaling pathway
Apoptosis - multiple species
Mitophagy - animal
Melanoma
Necroptosis
Axon guidance
C-type lectin receptor signaling pathway
0
5
10
15
-log10(qvalue)
C.
Proteoglycans in cancer
Cellular senescence
Pancreatic cancer
Adherens junction
Bladder cancer
AGE-RAGE signaling pathway in diabetic complications
Pathways in cancer
Human papillomavirus infection
Prostate cancer
Chronic myeloid leukemia
Hepatocellular carcinoma
Focal adhesion
Small cell lung cancer
0
2
4
6
-log10(qvalue)
D.
Proteoglycans in cancer
Focal adhesion
Cellular senescence
Protein processing in endoplasmic reticulum
Neurotrophin signaling pathway
FoxO signaling pathway
Pathways in cancer
Salmonella infection
Adherens junction
Prostate cancer
Ubiquitin mediated proteolysis
Small cell lung cancer
Human cytomegalovirus infection
Wnt signaling pathway
MAPK signaling pathway
ATP-dependent chromatin remodeling
Polycomb repressive complex
Autophagy - animal
Regulation of actin cytoskeleton
Shigellosis
mTOR signaling pathway
Colorectal cancer
p53 signaling pathway
Glioma
Endocrine resistance
Cell cycle
Kaposi sarcoma-associated herpesvirus infection
Human papillomavirus infection
Long-term potentiation
Lysine degradation
0
2
4
6
8
-log10(qvalue)

**Supplementary Fig. 3 Predictive modeling of post-vaccination miRNA expression influence on influenza vaccine-induced humoral immunity**

Modeling was performed as described in Statistical Analysis. The expressed post-vaccination CD4+ T cell and NK cell miRNAs predictive of influenza vaccine-induced memory B cell ELISPOT response are depicted in **A** and **B**, respectively. The CD4+ T cell miRNAs predictive of influenza vaccine-induced HAI antibody titer are depicted in **C**.

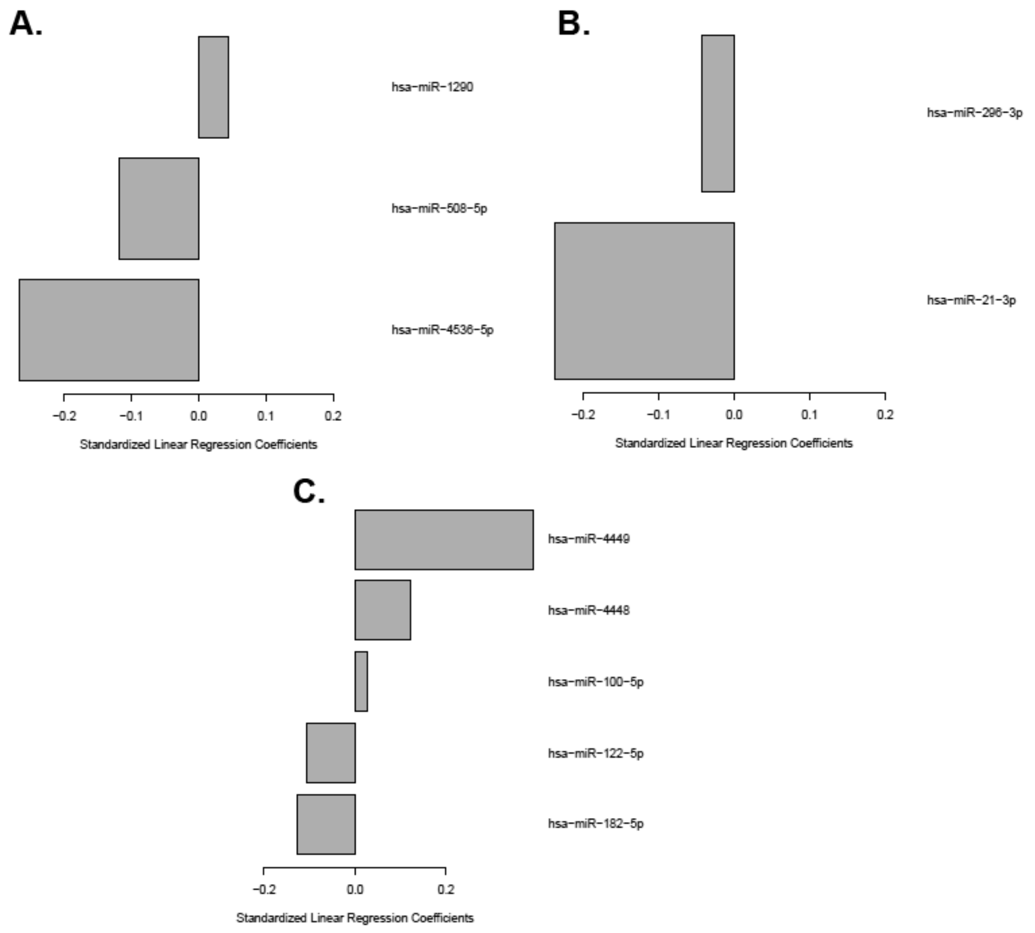

Supplementary Tables

**Supplementary Table 1. Highly expressed miRNAs in lymphocyte subsets**

| | *B cells* | | *CD4 T cells* | | *CD8 T cells* | | *NK cells* | |
|---|---|---|---|---|---|---|---|---|
| **miRNA** | **Median Counts** | **IQR** | **Median Counts** | **IQR** | **Median Counts** | **IQR** | **Median Counts** | **IQR** |
| hsa-let-7g-5p | 74158 | 52285, 116860 | 326077 | 161375, 434505 | 143762 | 87697, 252161 | 70904.5 | 24453.25, 117372.5 |
| hsa-miR-26a-5p | 52905 | 21383, 92863 | 136153 | 72309, 286052 | 71633 | 34696, 114194 | 39180 | 19538.25, 72709.25 |
| hsa-let-7i-5p | 47256 | 30947, 75218 | 83288 | 45034, 136022 | 41517 | 25149, 63880 | 28874 | 10700, 41428.5 |
| hsa-let-7f-5p | 42508 | 29542, 72432 | 83880 | 47010, 139908 | 53587 | 28161, 88174 | 59681 | 21127.5, 96118.5 |
| hsa-miR-92a-3p | 41636 | 23695, 53753 | 55656 | 35561, 84600 | 31787 | 22735, 50584 | 19963 | 8215.75, 26028.5 |
| hsa-let-7a-5p | 39957 | 28426, 72684 | 55012 | 36006, 77457 | 40766 | 24303, 65780 | 63405 | 23980.75, 99056.5 |
| hsa-miR-150-5p | 30878 | 17627, 48097 | 175344 | 123741, 305360 | 71002 | 47702, 100466 | 17583 | 5583, 38144.5 |
| hsa-miR-21-5p | 18938 | 9925, 31432 | 176401 | 74700, 394572 | 57906 | 28048, 130801 | 35599 | 17042.75, 65963.5 |
| hsa-miR-142-5p | 14613 | 5813, 27342 | 21578 | 9745, 46774 | 20266 | 7826, 29939 | 11882 | 5284.75, 21438 |
| hsa-miR-423-3p | 12170 | 8635, 17257 | 27531 | 18472, 37674 | 15502 | 11225, 25994 | 19340.5 | 6780.75, 27467.75 |
| hsa-miR-423-5p | 11421 | 7352, 15704 | 12175 | 8538, 17862 | 11603 | 7141, 19274 | 11597 | 6863, 18813.5 |
| hsa-miR-30d-5p | 9813 | 5735, 13267 | 21468 | 11036, 29545 | 12154 | 8403, 18898 | 10238.5 | 4030.5, 15021.75 |
| hsa-miR-155-5p | 9602 | 6959, 16732 | 9140 | 6119, 14734 | | | | |
| hsa-miR-181a-5p | 8657 | 3671, 15008 | 11202 | 4712, 19306 | 9380 | 4090, 16460 | 44730 | 20582.25, 80865.5 |
| hsa-let-7b-5p | 8046 | 5290, 13106 | 12250 | 7902, 18517 | 7758 | 5247, 11093 | 11241.5 | 7358, 18470.5 |
| hsa-miR-320a | 6220 | 3898, 7521 | | | 6831 | 4932, 9115 | 4989 | 2030.75, 6354 |
| hsa-miR-191-5p | 6056 | 3853, 10013 | | | 7641 | 3378, 9965 | 12164 | 6762, 23190.5 |
| hsa-miR-26b-5p | 5918 | 2778, 11138 | 12828 | 6889, 25415 | 10267 | 3909, 15520 | 9929.5 | 4396.5, 16903 |
| hsa-miR-101-3p | 4358 | 1457, 8333 | 11460 | 3787, 29872 | 6450 | 3596, 12073 | | |
| hsa-miR-140-3p | 3565 | 1807, 6860 | 9842 | 4581, 15583 | | | | |
| hsa-miR-146b-5p | | | 184810 | 71268, 287019 | 61250 | 32576, 125976 | 80884.5 | 34030.75, 173489.25 |
| hsa-miR-146a-5p | | | 23398 | 12198, 40894 | 10669 | 5570, 14783 | | |
| hsa-miR-181b-5p | | | | | | | 10392 | 4542, 17173 |
| hsa-miR-181a-2-3p | | | | | | | 5154 | 1793.5, 9938.5 |

An empty box indicates that the miRNA was filtered out of the corresponding cell subset hence there is no measurement.

| **Supplementary Table 2. miRNAs exhibiting expression changes following vaccination** | | | | | |
|---|---|---|---|---|---|
| **Cell subset** | **miRNA** | **Time** | **FC** | **p-value** | **q-value** |
| *$CD8^{+}$ T cell miRNAs* | | | | | |
| CD8 | hsa-miR-548f | Day 28 vs 0 | 0.48 | 0.00016 | 0.034 |
| CD8 | hsa-miR-196a-5p | Day 28 vs 0 | 0.54 | 0.00021 | 0.034 |
| CD8 | hsa-miR-9-3p | Day 28 vs 0 | 0.47 | 0.00047 | 0.046 |
| CD8 | hsa-miR-152 | Day 28 vs 0 | 0.66 | 0.00056 | 0.046 |
| CD8 | hsa-miR-9-5p | Day 28 vs 0 | 0.52 | 0.00080 | 0.052 |
| CD8 | hsa-miR-143-3p | Day 28 vs 0 | 0.45 | 0.0015 | 0.079 |
| *NK cell miRNAs* | | | | | |
| NK cell | hsa-miR-126-5p | Day 3 vs 0 | 0.38 | 0.0003 | 0.092 |
| NK cell | hsa-miR-148a-3p | Day 3 vs 0 | 0.56 | 0.0019 | 0.092 |
| NK cell | hsa-miR-4755-5p | Day 3 vs 0 | 0.50 | 0.0019 | 0.092 |
| NK cell | hsa-miR-339-5p | Day 3 vs 0 | 0.58 | 0.0022 | 0.092 |
| NK cell | hsa-miR-125b-5p | Day 3 vs 0 | 0.06 | 0.0023 | 0.092 |
| NK cell | hsa-miR-1291 | Day 3 vs 0 | 0.67 | 0.0024 | 0.092 |
| NK cell | hsa-miR-30a-3p | Day 3 vs 0 | 0.56 | 0.0035 | 0.092 |
| NK cell | hsa-miR-425-3p | Day 3 vs 0 | 0.59 | 0.0037 | 0.092 |
| NK cell | hsa-miR-500a-3p | Day 3 vs 0 | 0.58 | 0.0040 | 0.092 |
| NK cell | hsa-miR-203a | Day 3 vs 0 | 0.24 | 0.0041 | 0.092 |
| NK cell | hsa-miR-6501-5p | Day 3 vs 0 | 0.58 | 0.0046 | 0.092 |
| NK cell | hsa-miR-181c-5p | Day 3 vs 0 | 0.55 | 0.0051 | 0.092 |
| NK cell | hsa-miR-221-3p | Day 3 vs 0 | 0.61 | 0.0061 | 0.092 |
| NK cell | hsa-miR-1307-5p | Day 3 vs 0 | 0.55 | 0.0068 | 0.092 |
| NK cell | hsa-miR-550a-3-5p | Day 3 vs 0 | 0.56 | 0.007 | 0.092 |
| NK cell | hsa-miR-1275 | Day 3 vs 0 | 0.54 | 0.0074 | 0.092 |
| NK cell | hsa-miR-3158-3p | Day 3 vs 0 | 0.52 | 0.0075 | 0.092 |
| NK cell | hsa-miR-200a-3p | Day 3 vs 0 | 0.56 | 0.0078 | 0.092 |
| NK cell | hsa-miR-324-3p | Day 3 vs 0 | 0.50 | 0.0079 | 0.092 |
| NK cell | hsa-let-7i-5p | Day 3 vs 0 | 0.60 | 0.0089 | 0.092 |
| NK cell | hsa-miR-651 | Day 3 vs 0 | 0.54 | 0.0089 | 0.092 |
| NK cell | hsa-miR-20a-5p | Day 3 vs 0 | 0.59 | 0.0091 | 0.092 |
| NK cell | hsa-miR-10a-3p | Day 3 vs 0 | 0.58 | 0.0094 | 0.092 |
| NK cell | hsa-miR-194-5p | Day 3 vs 0 | 0.61 | 0.0100 | 0.092 |
| NK cell | hsa-miR-671-3p | Day 3 vs 0 | 0.63 | 0.010 | 0.092 |
| NK cell | hsa-miR-454-3p | Day 3 vs 0 | 0.64 | 0.010 | 0.092 |
| NK cell | hsa-miR-142-3p | Day 3 vs 0 | 0.58 | 0.011 | 0.092 |

| | | | | | |
|---|---|---|---|---|---|
| NK cell | hsa-miR-3613-5p | Day 3 vs 0 | 0.58 | 0.011 | 0.092 |
| NK cell | hsa-miR-590-3p | Day 3 vs 0 | 0.55 | 0.011 | 0.092 |
| NK cell | hsa-miR-1306-5p | Day 3 vs 0 | 0.68 | 0.011 | 0.092 |
| NK cell | hsa-miR-328 | Day 3 vs 0 | 0.68 | 0.012 | 0.092 |
| NK cell | hsa-miR-660-5p | Day 3 vs 0 | 0.60 | 0.012 | 0.092 |
| NK cell | hsa-miR-652-3p | Day 3 vs 0 | 0.58 | 0.012 | 0.092 |
| NK cell | hsa-miR-1292-5p | Day 3 vs 0 | 0.66 | 0.012 | 0.092 |
| NK cell | hsa-miR-15b-3p | Day 3 vs 0 | 0.63 | 0.012 | 0.092 |
| NK cell | hsa-miR-190b | Day 3 vs 0 | 0.70 | 0.012 | 0.092 |
| NK cell | hsa-miR-106b-3p | Day 3 vs 0 | 0.62 | 0.012 | 0.092 |
| NK cell | hsa-miR-17-5p | Day 3 vs 0 | 0.57 | 0.013 | 0.092 |
| NK cell | hsa-miR-140-5p | Day 3 vs 0 | 0.61 | 0.013 | 0.092 |
| NK cell | hsa-miR-93-5p | Day 3 vs 0 | 0.61 | 0.014 | 0.092 |
| NK cell | hsa-miR-200b-3p | Day 3 vs 0 | 0.61 | 0.014 | 0.092 |
| NK cell | hsa-miR-146a-5p | Day 3 vs 0 | 0.65 | 0.015 | 0.092 |
| NK cell | hsa-miR-99b-5p | Day 3 vs 0 | 0.17 | 0.015 | 0.092 |
| NK cell | hsa-miR-125a-5p | Day 3 vs 0 | 0.14 | 0.015 | 0.092 |
| NK cell | hsa-miR-27b-3p | Day 3 vs 0 | 0.58 | 0.015 | 0.092 |
| NK cell | hsa-miR-1294 | Day 3 vs 0 | 0.60 | 0.015 | 0.092 |
| NK cell | hsa-miR-32-3p | Day 3 vs 0 | 0.55 | 0.016 | 0.092 |
| NK cell | hsa-miR-107 | Day 3 vs 0 | 0.59 | 0.016 | 0.092 |
| NK cell | hsa-miR-151a-3p | Day 3 vs 0 | 0.58 | 0.016 | 0.092 |
| NK cell | hsa-miR-501-3p | Day 3 vs 0 | 0.63 | 0.016 | 0.092 |
| NK cell | hsa-miR-18a-3p | Day 3 vs 0 | 0.41 | 0.017 | 0.092 |
| NK cell | hsa-miR-625-5p | Day 3 vs 0 | 0.72 | 0.017 | 0.092 |
| NK cell | hsa-miR-98-5p | Day 3 vs 0 | 0.63 | 0.017 | 0.092 |
| NK cell | hsa-miR-671-5p | Day 3 vs 0 | 0.70 | 0.017 | 0.092 |
| NK cell | hsa-miR-3909 | Day 3 vs 0 | 0.55 | 0.018 | 0.092 |
| NK cell | hsa-miR-103a-3p | Day 3 vs 0 | 0.65 | 0.018 | 0.092 |
| NK cell | hsa-miR-1468 | Day 3 vs 0 | 0.62 | 0.018 | 0.092 |
| NK cell | hsa-miR-361-3p | Day 3 vs 0 | 0.62 | 0.018 | 0.092 |
| NK cell | hsa-miR-429 | Day 3 vs 0 | 0.51 | 0.018 | 0.092 |
| NK cell | hsa-miR-184 | Day 3 vs 0 | 0.63 | 0.019 | 0.092 |
| NK cell | hsa-miR-151a-5p | Day 3 vs 0 | 0.61 | 0.019 | 0.092 |
| NK cell | hsa-miR-3651 | Day 3 vs 0 | 0.66 | 0.019 | 0.092 |
| NK cell | hsa-miR-1271-5p | Day 3 vs 0 | 0.65 | 0.019 | 0.092 |
| NK cell | hsa-miR-345-5p | Day 3 vs 0 | 0.35 | 0.019 | 0.092 |
| NK cell | hsa-miR-25-3p | Day 3 vs 0 | 0.65 | 0.020 | 0.092 |
| NK cell | hsa-miR-30e-3p | Day 3 vs 0 | 0.64 | 0.020 | 0.092 |
| NK cell | hsa-let-7g-5p | Day 3 vs 0 | 0.61 | 0.021 | 0.092 |

| | | | | | |
|---|---|---|---|---|---|
| NK cell | hsa-miR-2277-5p | Day 3 vs 0 | 0.69 | 0.021 | 0.092 |
| NK cell | hsa-miR-642a-3p | Day 3 vs 0 | 0.66 | 0.021 | 0.092 |
| NK cell | hsa-miR-26b-5p | Day 3 vs 0 | 0.61 | 0.021 | 0.092 |
| NK cell | hsa-let-7c | Day 3 vs 0 | 0.65 | 0.022 | 0.092 |
| NK cell | hsa-miR-185-3p | Day 3 vs 0 | 0.68 | 0.022 | 0.092 |
| NK cell | hsa-miR-200c-3p | Day 3 vs 0 | 0.62 | 0.023 | 0.092 |
| NK cell | hsa-miR-132-5p | Day 3 vs 0 | 0.61 | 0.023 | 0.092 |
| NK cell | hsa-miR-589-5p | Day 3 vs 0 | 0.67 | 0.023 | 0.092 |
| NK cell | hsa-let-7e-5p | Day 3 vs 0 | 0.56 | 0.024 | 0.092 |
| NK cell | hsa-miR-29a-3p | Day 3 vs 0 | 0.62 | 0.024 | 0.092 |
| NK cell | hsa-miR-15a-5p | Day 3 vs 0 | 0.66 | 0.024 | 0.092 |
| NK cell | hsa-miR-181d | Day 3 vs 0 | 0.69 | 0.024 | 0.092 |
| NK cell | hsa-miR-339-3p | Day 3 vs 0 | 0.65 | 0.024 | 0.092 |
| NK cell | hsa-miR-874 | Day 3 vs 0 | 0.70 | 0.024 | 0.092 |
| NK cell | hsa-miR-484 | Day 3 vs 0 | 0.66 | 0.024 | 0.092 |
| NK cell | hsa-miR-30d-3p | Day 3 vs 0 | 0.56 | 0.024 | 0.092 |
| NK cell | hsa-miR-532-5p | Day 3 vs 0 | 0.64 | 0.024 | 0.092 |
| NK cell | hsa-let-7a-5p | Day 3 vs 0 | 0.67 | 0.025 | 0.092 |
| NK cell | hsa-miR-4521 | Day 3 vs 0 | 0.66 | 0.025 | 0.092 |
| NK cell | hsa-let-7f-5p | Day 3 vs 0 | 0.63 | 0.025 | 0.092 |
| NK cell | hsa-miR-106b-5p | Day 3 vs 0 | 0.65 | 0.025 | 0.092 |
| NK cell | hsa-let-7d-5p | Day 3 vs 0 | 0.67 | 0.026 | 0.092 |
| NK cell | hsa-miR-629-5p | Day 3 vs 0 | 0.63 | 0.026 | 0.092 |
| NK cell | hsa-miR-378i | Day 3 vs 0 | 0.61 | 0.026 | 0.092 |
| NK cell | hsa-miR-941 | Day 3 vs 0 | 0.65 | 0.028 | 0.093 |
| NK cell | hsa-miR-877-5p | Day 3 vs 0 | 0.57 | 0.028 | 0.093 |
| NK cell | hsa-miR-222-3p | Day 3 vs 0 | 0.67 | 0.029 | 0.093 |
| NK cell | hsa-miR-23b-3p | Day 3 vs 0 | 0.60 | 0.029 | 0.093 |
| NK cell | hsa-miR-502-3p | Day 3 vs 0 | 0.65 | 0.029 | 0.093 |
| NK cell | hsa-miR-1307-3p | Day 3 vs 0 | 0.72 | 0.029 | 0.093 |
| NK cell | hsa-miR-148b-3p | Day 3 vs 0 | 0.66 | 0.029 | 0.093 |
| NK cell | hsa-miR-335-3p | Day 3 vs 0 | 0.53 | 0.030 | 0.093 |
| NK cell | hsa-miR-409-3p | Day 3 vs 0 | 0.57 | 0.030 | 0.093 |
| NK cell | hsa-miR-24-3p | Day 3 vs 0 | 0.68 | 0.030 | 0.093 |
| NK cell | hsa-miR-550a-5p | Day 3 vs 0 | 0.58 | 0.030 | 0.093 |
| NK cell | hsa-miR-21-3p | Day 3 vs 0 | 0.33 | 0.031 | 0.093 |
| NK cell | hsa-miR-152 | Day 3 vs 0 | 0.67 | 0.031 | 0.093 |
| NK cell | hsa-miR-210 | Day 3 vs 0 | 0.31 | 0.031 | 0.093 |
| NK cell | hsa-miR-15b-5p | Day 3 vs 0 | 0.58 | 0.031 | 0.093 |
| NK cell | hsa-miR-505-3p | Day 3 vs 0 | 0.65 | 0.031 | 0.093 |

| | | | | | |
|---|---|---|---|---|---|
| NK cell | hsa-miR-29b-3p | Day 3 vs 0 | 0.65 | 0.031 | 0.093 |
| NK cell | hsa-miR-574-5p | Day 3 vs 0 | 0.49 | 0.031 | 0.093 |
| NK cell | hsa-miR-664b-5p | Day 3 vs 0 | 0.68 | 0.032 | 0.093 |
| NK cell | hsa-miR-27b-5p | Day 3 vs 0 | 0.60 | 0.032 | 0.093 |
| NK cell | hsa-miR-1287 | Day 3 vs 0 | 0.67 | 0.033 | 0.094 |
| NK cell | hsa-miR-30c-5p | Day 3 vs 0 | 0.66 | 0.034 | 0.094 |
| NK cell | hsa-miR-212-5p | Day 3 vs 0 | 0.66 | 0.034 | 0.094 |
| NK cell | hsa-miR-186-5p | Day 3 vs 0 | 0.66 | 0.034 | 0.094 |
| NK cell | hsa-let-7d-3p | Day 3 vs 0 | 0.68 | 0.034 | 0.094 |
| NK cell | hsa-miR-324-5p | Day 3 vs 0 | 0.60 | 0.035 | 0.094 |
| NK cell | hsa-miR-185-5p | Day 3 vs 0 | 0.65 | 0.035 | 0.094 |
| NK cell | hsa-miR-92b-3p | Day 3 vs 0 | 0.21 | 0.035 | 0.094 |
| NK cell | hsa-miR-374a-3p | Day 3 vs 0 | 0.59 | 0.035 | 0.094 |
| NK cell | hsa-miR-296-3p | Day 3 vs 0 | 0.44 | 0.035 | 0.094 |
| NK cell | hsa-miR-330-5p | Day 3 vs 0 | 0.67 | 0.036 | 0.094 |
| NK cell | hsa-miR-22-3p | Day 3 vs 0 | 0.65 | 0.036 | 0.094 |
| NK cell | hsa-miR-1248 | Day 3 vs 0 | 0.70 | 0.036 | 0.095 |
| NK cell | hsa-miR-106a-5p | Day 3 vs 0 | 0.69 | 0.037 | 0.095 |
| NK cell | hsa-miR-29c-3p | Day 3 vs 0 | 0.64 | 0.037 | 0.095 |
| NK cell | hsa-miR-628-3p | Day 3 vs 0 | 0.67 | 0.038 | 0.096 |
| NK cell | hsa-miR-1908 | Day 3 vs 0 | 0.68 | 0.038 | 0.096 |
| NK cell | hsa-miR-92a-3p | Day 3 vs 0 | 0.69 | 0.039 | 0.098 |
| NK cell | hsa-miR-30b-3p | Day 3 vs 0 | 0.70 | 0.039 | 0.098 |
| NK cell | hsa-miR-873-3p | Day 3 vs 0 | 0.69 | 0.040 | 0.098 |
| NK cell | hsa-miR-22-5p | Day 3 vs 0 | 0.65 | 0.040 | 0.098 |
| NK cell | hsa-miR-128 | Day 3 vs 0 | 0.65 | 0.040 | 0.098 |
| NK cell | hsa-miR-1268b | Day 3 vs 0 | 0.61 | 0.041 | 0.098 |
| NK cell | hsa-miR-421 | Day 3 vs 0 | 0.71 | 0.041 | 0.098 |
| NK cell | hsa-miR-769-5p | Day 3 vs 0 | 0.67 | 0.042 | 0.098 |
| NK cell | hsa-miR-1268a | Day 3 vs 0 | 0.65 | 0.042 | 0.098 |
| NK cell | hsa-miR-5701 | Day 3 vs 0 | 0.74 | 0.042 | 0.098 |
| NK cell | hsa-miR-942 | Day 3 vs 0 | 0.72 | 0.042 | 0.098 |
| NK cell | hsa-miR-1273g-3p | Day 3 vs 0 | 0.67 | 0.043 | 0.099 |
| NK cell | hsa-miR-362-5p | Day 3 vs 0 | 0.63 | 0.043 | 0.099 |
| NK cell | hsa-miR-374a-5p | Day 3 vs 0 | 0.66 | 0.043 | 0.099 |

**Supplementary Table 3. Baseline miRNAs correlated with HAI, VNA and B cell ELISPOT response**

| miRNA | Time-point | B cell Correlation | B cell p-value | CD4 Correlation | CD4 p-value | CD8 Correlation | CD8 p-value | NK Correlation | NK p-value |
|---|---|---|---|---|---|---|---|---|---|
| **Correlations with HAI response** | | | | | | | | | |
| hsa-miR-331-3p | Day 0 | **0.50** | **0.00076** | **0.41** | **0.0077** | 0.42 | 0.016 | **0.44** | **0.0089** |
| hsa-miR-4448 | Day 0 | **0.49** | **0.0010** | 0.35 | 0.025 | 0.39 | 0.026 | **0.44** | **0.0094** |
| hsa-miR-766-3p | Day 0 | **0.44** | **0.0045** | 0.15 | 0.35 | 0.01 | 0.963 | 0.06 | 0.72 |
| hsa-miR-342-3p | Day 0 | **0.43** | **0.0047** | 0.32 | 0.041 | 0.32 | 0.070 | 0.40 | 0.018 |
| hsa-miR-4792 | Day 0 | **0.43** | **0.0048** | 0.14 | 0.39 | 0.40 | 0.022 | 0.32 | 0.065 |
| hsa-miR-15b-3p | Day 0 | **0.42** | **0.0058** | 0.33 | 0.033 | 0.43 | 0.012 | 0.41 | 0.016 |
| hsa-miR-1292-5p | Day 0 | **-0.41** | **0.0074** | -0.17 | 0.28 | -0.31 | 0.079 | -0.27 | 0.13 |
| hsa-miR-3916 | Day 0 | **0.41** | **0.0078** | -0.17 | 0.30 | -0.14 | 0.422 | 0.23 | 0.18 |
| hsa-miR-18a-5p | Day 0 | **0.41** | **0.0081** | -0.01 | 0.97 | 0.36 | 0.042 | | |
| hsa-miR-660-5p | Day 0 | 0.35 | 0.026 | 0.37 | 0.017 | **0.46** | **0.0076** | 0.21 | 0.24 |
| hsa-miR-28-3p | Day 0 | -0.33 | 0.033 | -0.17 | 0.29 | **-0.61** | **0.00014** | **-0.47** | **0.0047** |
| hsa-miR-107 | Day 0 | 0.33 | 0.034 | **0.50** | **0.00095** | 0.42 | 0.015 | 0.38 | 0.025 |
| hsa-miR-122-5p | Day 0 | -0.33 | 0.035 | **-0.43** | **0.0053** | **-0.48** | **0.0047** | -0.39 | 0.024 |
| hsa-miR-27a-5p | Day 0 | -0.32 | 0.042 | -0.33 | 0.033 | **-0.52** | **0.0020** | -0.32 | 0.069 |
| hsa-miR-150-3p | Day 0 | -0.31 | 0.049 | -0.31 | 0.048 | **-0.49** | **0.0035** | **-0.59** | **0.00026** |
| hsa-miR-199a-3p | Day 0 | 0.29 | 0.064 | 0.34 | 0.029 | **0.64** | **0.00007** | **0.58** | **0.00031** |
| hsa-miR-181b-5p | Day 0 | 0.29 | 0.067 | 0.35 | 0.024 | 0.11 | 0.557 | **0.46** | **0.0060** |
| hsa-miR-2110 | Day 0 | -0.29 | 0.069 | -0.19 | 0.23 | **-0.51** | **0.0026** | **-0.55** | **0.00072** |
| hsa-miR-4449 | Day 0 | 0.29 | 0.071 | **0.43** | **0.0046** | 0.29 | 0.106 | | |
| hsa-miR-3934-5p | Day 0 | -0.29 | 0.071 | 0.14 | 0.37 | **-0.53** | **0.0017** | -0.33 | 0.054 |
| hsa-miR-199b-3p | Day 0 | 0.28 | 0.079 | 0.35 | 0.027 | **0.65** | **0.000047** | **0.53** | **0.0011** |
| hsa-miR-4443 | Day 0 | -0.27 | 0.094 | -0.16 | 0.32 | **-0.47** | **0.0054** | **-0.56** | **0.00062** |
| hsa-miR-30a-5p | Day 0 | 0.24 | 0.13 | 0.07 | 0.67 | **0.45** | **0.0092** | 0.17 | 0.34 |
| hsa-miR-423-5p | Day 0 | -0.24 | 0.13 | -0.29 | 0.064 | -0.43 | 0.013 | **-0.51** | **0.0023** |
| hsa-miR-181a-5p | Day 0 | 0.23 | 0.14 | 0.35 | 0.023 | 0.18 | 0.322 | **0.49** | **0.0035** |
| hsa-miR-199b-5p | Day 0 | 0.19 | 0.24 | 0.23 | 0.14 | **0.57** | **0.00052** | | |
| hsa-miR-1255a | Day 0 | -0.17 | 0.30 | -0.22 | 0.17 | **-0.58** | **0.00041** | -0.23 | 0.20 |
| hsa-miR-330-3p | Day 0 | -0.14 | 0.38 | -0.16 | 0.32 | **-0.53** | **0.0013** | -0.24 | 0.18 |
| hsa-miR-17-3p | Day 0 | 0.14 | 0.40 | 0.22 | 0.17 | **0.58** | **0.00045** | | |
| hsa-miR-744-5p | Day 0 | -0.11 | 0.49 | -0.19 | 0.24 | **-0.49** | **0.0042** | -0.23 | 0.19 |
| hsa-miR-629-5p | Day 0 | -0.11 | 0.51 | -0.35 | 0.024 | **-0.54** | **0.0012** | -0.42 | 0.014 |
| hsa-miR-589-5p | Day 0 | -0.08 | 0.60 | -0.07 | 0.68 | **-0.45** | **0.0080** | -0.15 | 0.41 |
| hsa-miR-423-3p | Day 0 | -0.06 | 0.71 | 0.08 | 0.62 | -0.32 | 0.069 | **-0.45** | **0.0083** |
| hsa-miR-505-3p | Day 0 | 0.05 | 0.75 | 0.32 | 0.043 | 0.14 | 0.425 | **0.48** | **0.0040** |
| hsa-miR-155-5p | Day 0 | 0.01 | 0.95 | -0.16 | 0.33 | -0.22 | 0.209 | **-0.50** | **0.0028** |

| | | | | | | | | | |
|---|---|---|---|---|---|---|---|---|---|
| hsa-miR-486-3p | Day 0 | 0.01 | 0.96 | **-0.40** | **0.0097** | -0.02 | 0.908 | | |
| hsa-miR-3179 | Day 0 | | | **-0.40** | **0.0091** | -0.32 | 0.070 | -0.20 | 0.26 |
| hsa-miR-3199 | Day 0 | | | **0.40** | **0.0095** | | | | |
| hsa-miR-4999-5p | Day 0 | | | -0.09 | 0.58 | **-0.46** | **0.0072** | | |
| **Correlations with VNA response** | | | | | | | | | |
| hsa-miR-331-3p | Day 0 | **0.49** | **0.0011** | **0.44** | **0.0045** | 0.43 | 0.013 | 0.42 | 0.013 |
| hsa-miR-4792 | Day 0 | **0.48** | **0.0016** | 0.21 | 0.18 | **0.51** | **0.0022** | 0.42 | 0.012 |
| hsa-miR-1292-5p | Day 0 | **-0.47** | **0.0018** | -0.18 | 0.25 | -0.34 | 0.052 | -0.18 | 0.31 |
| hsa-miR-1248 | Day 0 | **0.47** | **0.0018** | 0.22 | 0.17 | 0.25 | 0.16 | 0.34 | 0.051 |
| hsa-miR-3916 | Day 0 | **0.47** | **0.002** | -0.12 | 0.45 | -0.09 | 0.61 | 0.27 | 0.12 |
| hsa-miR-342-3p | Day 0 | **0.44** | **0.0044** | **0.40** | **0.0087** | **0.46** | **0.0076** | 0.42 | 0.013 |
| hsa-miR-4491 | Day 0 | **-0.43** | **0.0049** | | | | | | |
| hsa-miR-191-5p | Day 0 | **0.42** | **0.0068** | 0.23 | 0.16 | 0.09 | 0.61 | 0.32 | 0.062 |
| hsa-miR-4448 | Day 0 | **0.41** | **0.0079** | 0.27 | 0.087 | 0.24 | 0.18 | 0.42 | 0.014 |
| hsa-miR-484 | Day 0 | **0.41** | **0.0083** | 0.28 | 0.078 | 0.32 | 0.071 | 0.14 | 0.44 |
| hsa-miR-150-5p | Day 0 | 0.37 | 0.016 | 0.40 | 0.0087 | 0.42 | 0.015 | 0.23 | 0.19 |
| hsa-miR-3934-5p | Day 0 | -0.36 | 0.02 | 0.01 | 0.94 | **-0.46** | **0.0066** | -0.29 | 0.092 |
| hsa-miR-27a-5p | Day 0 | -0.34 | 0.028 | **-0.46** | **0.0026** | **-0.61** | **0.00015** | **-0.48** | **0.0037** |
| hsa-miR-2110 | Day 0 | -0.34 | 0.031 | -0.26 | 0.1 | **-0.55** | **0.00087** | **-0.52** | **0.0017** |
| hsa-miR-4449 | Day 0 | 0.31 | 0.05 | **0.42** | **0.0068** | 0.23 | 0.21 | | |
| hsa-miR-150-3p | Day 0 | -0.29 | 0.065 | **-0.42** | **0.0063** | **-0.54** | **0.0012** | **-0.59** | **0.00025** |
| hsa-miR-199b-5p | Day 0 | 0.29 | 0.065 | 0.29 | 0.068 | **0.64** | **0.000068** | | |
| hsa-miR-28-3p | Day 0 | -0.29 | 0.068 | -0.31 | 0.051 | **-0.64** | **0.000064** | **-0.52** | **0.0014** |
| hsa-miR-660-5p | Day 0 | 0.29 | 0.068 | **0.44** | **0.0041** | **0.44** | **0.0096** | 0.16 | 0.36 |
| hsa-miR-1255a | Day 0 | -0.28 | 0.081 | -0.25 | 0.11 | **-0.65** | **0.000036** | -0.31 | 0.075 |
| hsa-let-7c | Day 0 | -0.26 | 0.099 | -0.04 | 0.78 | -0.32 | 0.068 | **-0.44** | **0.0087** |
| hsa-miR-423-5p | Day 0 | -0.25 | 0.11 | -0.33 | 0.033 | **-0.49** | **0.0039** | **-0.47** | **0.0048** |
| hsa-miR-29b-2-5p | Day 0 | 0.25 | 0.11 | 0.19 | 0.25 | **0.46** | **0.007** | 0.30 | 0.079 |
| hsa-miR-4443 | Day 0 | -0.25 | 0.12 | -0.24 | 0.13 | **-0.54** | **0.0012** | **-0.54** | **0.0011** |
| hsa-miR-122-5p | Day 0 | -0.25 | 0.12 | -0.38 | 0.013 | **-0.52** | **0.002** | -0.19 | 0.28 |
| hsa-miR-199a-3p | Day 0 | 0.24 | 0.13 | 0.28 | 0.078 | **0.58** | **0.0004** | **0.50** | **0.0026** |
| hsa-miR-125b-5p | Day 0 | 0.23 | 0.15 | **0.49** | **0.0011** | 0.34 | 0.054 | 0.16 | 0.36 |
| hsa-miR-107 | Day 0 | 0.23 | 0.15 | **0.40** | **0.0088** | 0.44 | 0.011 | 0.22 | 0.21 |
| hsa-miR-99a-5p | Day 0 | 0.23 | 0.16 | **0.41** | **0.0083** | **0.48** | **0.0049** | 0.33 | 0.054 |
| hsa-miR-199b-3p | Day 0 | 0.22 | 0.17 | 0.28 | 0.078 | **0.59** | **0.0003** | **0.53** | **0.0011** |
| hsa-let-7i-5p | Day 0 | -0.20 | 0.22 | -0.30 | 0.058 | **-0.48** | **0.0046** | -0.38 | 0.026 |
| hsa-miR-766-5p | Day 0 | -0.18 | 0.25 | -0.15 | 0.33 | **-0.46** | **0.0075** | -0.21 | 0.24 |
| hsa-miR-629-5p | Day 0 | -0.17 | 0.3 | **-0.40** | **0.0095** | **-0.62** | **0.0001** | -0.41 | 0.017 |
| hsa-miR-4446-3p | Day 0 | -0.16 | 0.31 | -0.38 | 0.015 | **-0.48** | **0.0045** | -0.33 | 0.054 |
| hsa-let-7a-5p | Day 0 | -0.16 | 0.32 | -0.21 | 0.19 | -0.34 | 0.054 | **-0.52** | **0.0018** |

| | | | | | | | | | |
|---|---|---|---|---|---|---|---|---|---|
| hsa-miR-181a-5p | Day 0 | 0.14 | 0.37 | **0.42** | **0.0068** | 0.29 | 0.1 | **0.49** | **0.0035** |
| hsa-miR-330-3p | Day 0 | -0.14 | 0.37 | -0.22 | 0.16 | -0.61 | 0.00016 | -0.26 | 0.13 |
| hsa-miR-378a-5p | Day 0 | 0.14 | 0.38 | 0.12 | 0.44 | 0.50 | 0.0033 | | |
| hsa-miR-146b-5p | Day 0 | -0.14 | 0.39 | -0.09 | 0.59 | 0.08 | 0.65 | **-0.50** | **0.0028** |
| hsa-miR-744-5p | Day 0 | -0.13 | 0.43 | -0.27 | 0.086 | **-0.52** | **0.0019** | -0.27 | 0.13 |
| hsa-miR-200b-5p | Day 0 | -0.11 | 0.48 | | | | | **-0.50** | **0.0025** |
| hsa-miR-423-3p | Day 0 | -0.08 | 0.6 | 0.03 | 0.85 | -0.27 | 0.13 | **-0.48** | **0.0044** |
| hsa-miR-505-3p | Day 0 | 0.08 | 0.63 | 0.34 | 0.031 | 0.06 | 0.74 | **0.48** | **0.0039** |
| hsa-let-7d-3p | Day 0 | 0.06 | 0.7 | -0.04 | 0.79 | -0.21 | 0.24 | **-0.52** | **0.0016** |
| hsa-miR-3064-5p | Day 0 | -0.06 | 0.72 | -0.37 | 0.017 | -0.37 | 0.036 | **-0.48** | **0.0037** |
| hsa-miR-486-3p | Day 0 | 0.05 | 0.76 | **-0.46** | **0.0026** | -0.15 | 0.4 | | |
| hsa-miR-155-5p | Day 0 | 0.05 | 0.77 | -0.19 | 0.25 | -0.25 | 0.16 | **-0.45** | **0.0073** |
| hsa-miR-589-5p | Day 0 | -0.04 | 0.79 | -0.11 | 0.48 | **-0.45** | **0.0086** | -0.13 | 0.46 |
| hsa-miR-17-3p | Day 0 | 0.02 | 0.9 | 0.22 | 0.17 | **0.51** | **0.0027** | | |
| hsa-miR-3177-3p | Day 0 | | | 0.16 | 0.32 | **-0.47** | **0.006** | -0.36 | 0.037 |
| hsa-miR-3179 | Day 0 | | | -0.39 | 0.011 | **-0.46** | **0.0069** | -0.09 | 0.61 |
| hsa-miR-324-5p | Day 0 | | | 0.24 | 0.14 | **0.47** | **0.0057** | 0.02 | 0.91 |
| hsa-miR-3928 | Day 0 | | | -0.28 | 0.074 | | | **-0.44** | **0.009** |
| hsa-miR-4661-5p | Day 0 | | | -0.10 | 0.55 | **-0.50** | **0.0031** | | |
| hsa-miR-6130 | Day 0 | | | -0.17 | 0.28 | | | **-0.46** | **0.0065** |
| **Correlations with B cell ELISPOT response** | | | | | | | | | |
| hsa-miR-150-3p | Day 0 | **-0.53** | **0.00038** | -0.38 | 0.015 | -0.28 | 0.11 | -0.3 | 0.083 |
| hsa-miR-652-3p | Day 0 | **0.53** | **0.0004** | 0.21 | 0.18 | 0.24 | 0.18 | 0.22 | 0.2 |
| hsa-let-7d-3p | Day 0 | **-0.49** | **0.0012** | **-0.41** | **0.0079** | -0.41 | 0.018 | -0.12 | 0.5 |
| hsa-let-7c | Day 0 | **-0.47** | **0.0022** | -0.33 | 0.034 | **-0.52** | **0.0018** | -0.39 | 0.023 |
| hsa-miR-1275 | Day 0 | **-0.47** | **0.0022** | -0.23 | 0.15 | 0.01 | 0.94 | -0.01 | 0.96 |
| hsa-miR-629-5p | Day 0 | **-0.46** | **0.0025** | -0.27 | 0.091 | -0.35 | 0.048 | -0.07 | 0.7 |
| hsa-miR-30a-5p | Day 0 | **0.46** | **0.0026** | 0.15 | 0.35 | -0.16 | 0.37 | -0.14 | 0.44 |
| hsa-miR-1268a | Day 0 | **-0.45** | **0.0028** | -0.17 | 0.29 | -0.43 | 0.012 | -0.17 | 0.33 |
| hsa-miR-20b-5p | Day 0 | **0.45** | **0.0028** | 0.23 | 0.15 | -0.1 | 0.59 | 0.02 | 0.93 |
| hsa-let-7b-5p | Day 0 | **-0.45** | **0.0029** | -0.26 | 0.1 | -0.32 | 0.074 | -0.37 | 0.03 |
| hsa-miR-25-5p | Day 0 | **-0.44** | **0.0037** | -0.34 | 0.028 | -0.3 | 0.089 | -0.39 | 0.021 |
| hsa-miR-4443 | Day 0 | **-0.44** | **0.0044** | -0.33 | 0.037 | -0.38 | 0.03 | -0.32 | 0.068 |
| hsa-miR-1268b | Day 0 | **-0.43** | **0.0046** | -0.2 | 0.2 | -0.44 | 0.011 | -0.16 | 0.37 |
| hsa-miR-103a-3p | Day 0 | **0.43** | **0.0049** | 0.16 | 0.31 | 0.11 | 0.55 | -0.04 | 0.81 |
| hsa-miR-223-3p | Day 0 | **0.43** | **0.0051** | 0.16 | 0.31 | 0.13 | 0.46 | 0.16 | 0.36 |
| hsa-miR-15b-5p | Day 0 | **0.43** | **0.0052** | 0.33 | 0.038 | 0.17 | 0.36 | 0.07 | 0.69 |
| hsa-miR-17-5p | Day 0 | **0.42** | **0.0064** | 0.17 | 0.3 | 0.03 | 0.89 | -0.03 | 0.88 |
| hsa-miR-5196-3p | Day 0 | **-0.41** | **0.0072** | | | | | | |
| hsa-miR-92a-1-5p | Day 0 | **-0.41** | **0.0078** | -0.23 | 0.15 | -0.35 | 0.048 | -0.33 | 0.059 |

| | | | | | | | | | |
|---|---|---|---|---|---|---|---|---|---|
| hsa-miR-106b-3p | Day 0 | **0.4** | **0.009** | 0.02 | 0.91 | -0.12 | 0.49 | 0.22 | 0.21 |
| hsa-miR-664a-5p | Day 0 | **-0.4** | **0.0097** | -0.34 | 0.031 | -0.32 | 0.067 | -0.12 | 0.51 |
| hsa-miR-23b-3p | Day 0 | 0.35 | 0.025 | **0.44** | **0.0043** | 0.27 | 0.12 | 0.12 | 0.51 |
| hsa-miR-374b-5p | Day 0 | 0.35 | 0.025 | **0.51** | **0.00075** | 0.28 | 0.12 | 0.13 | 0.45 |
| hsa-miR-29b-1-5p | Day 0 | -0.33 | 0.034 | 0.07 | 0.68 | **0.48** | **0.005** | | |
| hsa-miR-425-5p | Day 0 | 0.33 | 0.034 | **0.5** | **0.00093** | 0.19 | 0.29 | 0.27 | 0.13 |
| hsa-miR-1468 | Day 0 | -0.3 | 0.056 | -0.21 | 0.18 | **-0.45** | **0.0094** | 0.24 | 0.18 |
| hsa-miR-342-5p | Day 0 | -0.28 | 0.071 | **-0.4** | **0.0099** | -0.28 | 0.12 | -0.22 | 0.21 |
| hsa-miR-23a-3p | Day 0 | 0.26 | 0.095 | **0.43** | **0.005** | 0.21 | 0.23 | 0.08 | 0.67 |
| hsa-miR-18a-5p | Day 0 | 0.25 | 0.11 | **0.46** | **0.0024** | 0.08 | 0.65 | | |
| hsa-miR-3173-5p | Day 0 | -0.23 | 0.14 | **-0.41** | **0.0077** | -0.2 | 0.26 | -0.17 | 0.34 |
| hsa-miR-671-3p | Day 0 | -0.23 | 0.16 | **-0.42** | **0.0063** | -0.31 | 0.083 | 0.11 | 0.53 |
| hsa-miR-30b-5p | Day 0 | 0.22 | 0.17 | **0.54** | **0.0003** | 0.16 | 0.36 | 0.12 | 0.51 |
| hsa-miR-1248 | Day 0 | 0.15 | 0.34 | 0.07 | 0.64 | **0.53** | **0.0015** | 0.37 | 0.033 |
| hsa-miR-128 | Day 0 | -0.13 | 0.42 | -0.08 | 0.61 | **-0.45** | **0.0087** | 0.04 | 0.84 |
| hsa-miR-19b-3p | Day 0 | 0.07 | 0.65 | **0.4** | **0.0095** | 0 | 1 | 0.23 | 0.19 |
| hsa-miR-29c-3p | Day 0 | -0.06 | 0.71 | **0.45** | **0.0035** | 0.17 | 0.34 | -0.06 | 0.73 |
| hsa-miR-1273g-3p | Day 0 | -0.04 | 0.79 | 0.14 | 0.39 | **0.51** | **0.0022** | 0.01 | 0.97 |
| hsa-miR-501-3p | Day 0 | 0.03 | 0.88 | **-0.42** | **0.0058** | -0.16 | 0.36 | -0.21 | 0.23 |
| hsa-miR-200b-3p | Day 0 | -0.02 | 0.89 | -0.16 | 0.32 | **-0.5** | **0.0028** | -0.02 | 0.9 |

An empty box indicates that the miRNA was filtered out of the corresponding cell subset hence there is no measurement.
